# The impact of COVID-19 non-pharmaceutical interventions on the future dynamics of endemic infections

**DOI:** 10.1101/2020.06.22.20137588

**Authors:** Rachel E. Baker, Sang Woo Park, Wenchang Yang, Gabriel A. Vecchi, C. Jessica E. Metcalf, Bryan T. Grenfell

## Abstract

Non-pharmaceutical interventions (NPIs) have been employed to reduce the transmission of SARS-CoV-2, yet these measures are already having similar effects on other directly-transmitted, endemic diseases. Disruptions to the seasonal transmission patterns of these diseases may have consequences for the timing and severity of future outbreaks. Here we consider the implications of SARS-CoV-2 NPIs for two endemic infections circulating in the United States of America (USA): respiratory syncytial virus (RSV) and seasonal influenza. Using laboratory surveillance data from 2020, we estimate that RSV transmission declined by at least 20% in the USA at the start of the NPI period. We simulate future trajectories of both RSV and influenza, using an epidemic model. As susceptibility increases over the NPI period, we find that substantial outbreaks of RSV may occur in future years, with peak outbreaks likely occurring in the winter of 2021-2022. Results for influenza broadly echo this picture, but are more uncertain; future outbreaks are likely dependent on the transmissibility and evolutionary dynamics of circulating strains.

## Main Text

Non-pharmaceutical interventions (NPIs) have proved effective in reducing the spread of SARS-CoV-2 in many contexts [1, 2, 3, 4, 5]. Policy measures including social-distancing, school closures, travel restrictions and the use of masks in public spaces have been implemented to reduce the transmission of the virus. In addition to SARS-CoV-2, NPIs may also reduce the transmission of other directly-transmitted, respiratory infections [6, 7]. Understanding the possible influence of a SARS-CoV-2 NPI period on the incidence of these infections remains a key question for the broader public health impact of the pandemic. Furthermore, the implications of relaxing NPIs for future outbreaks of these other infections have not been fully considered.

Many endemic, directly-transmitted, respiratory infections exhibit distinct seasonal and longer term cycles in incidence [8, 9, 10]. While climate may drive the seasonality of these diseases in some cases [11, 12, 13, 14], other directly-transmitted infections, such as measles, are driven primarily by seasonal cycles of population aggregation such as the timing of school semesters [15, 16]. Secular changes in susceptible recruitment, for instance due to vaccination campaigns or declines in birth rates, can disrupt long-run patterns of infection dynamics [17, 18]. Similarly, human movement via either displacement or migration, has also been shown to alter patterns of infection [19]. While there has been less work to identify the polymicrobial implications of non-pharmaceutical control measures, evidence from the 1918 influenza pandemic suggests that NPIs may have reduced measles transmission by 38% [20].

Two important directly-transmitted, viral respiratory diseases circulating in the USA population are seasonal influenza and respiratory syncytial virus (RSV). Seasonal influenza accounts for significant annual mortality, with the ongoing evolution of the virus’ antigenic sites leading to evasion of the host immune system [21, 22]. Epidemics of seasonal influenza at higher latitudes are driven largely by variations in climate [12, 13]. While there is some evidence of herd immunity, a complex interaction between alternating sub-types and antigenic drift determines year-to-year variation in susceptibility and corresponding outbreak size [23, 10].

RSV causes lower respiratory tract infections in young infants, and contributes to approximately 5% of under-five deaths globally [24], with no vaccine currently available. Previous models show RSV epidemics exhibit limit cycle behavior, tuned by climate-driven seasonality (*Methods*) [25, 11]. In most regions in the USA, RSV and influenza exhibit peak incidence in the winter months, coinciding with cold, dry climatic conditions [13, 11].

Here we consider the impact of non-pharmaceutical control measures on the incidence of these two respiratory infections. We focus primarily on RSV, with the simpler limit cycle dynamics presenting an opportunity to probe interactions with NPIs. We first evaluate the influence of control measures targeting SARS-CoV-2 using influenza and RSV surveillance data. Since changes to physician visits for both viruses could be driven by behavioral responses to control measures, we look at the percent positive tests for both viruses as reported from laboratory surveillance data.

Fig 1 shows the percent positive tests for RSV (Fig 1a) and influenza (Fig 1b) for 2019-2020 (highlighted) and four preceding years, for four states (RSV data with at least two years of observations were not available for other states). A national emergency in response to theCOVID-19 pandemic was declared on March 13th 2020 in the US, shown with the dashed line. Following the declaration, many states put in place control measures to limit the spread of SARS-CoV-2. Despite the declaration occurring after the typical seasonal peak in cases, a decline in prevalence is observed beyond mean seasonal levels. In Florida, where RSV cases tend to persist throughout the year [11, 25], observed RSV prevalence is reduced to near zero in March 2020. A similar pattern is visible in Hawaii for influenza, where cases are normally persistent. In Fig 1c we show the 2019-2020 change in percentage positive influenza tests relative to weekly mean over the previous four seasons. The 2019-2020 influenza season appears to have been more severe than average, with a relative increase in prevalence prior to March 2020 possibly driven by increased circulation of influenza subtype B (Fig. S2). However, following the declaration of emergency, declines to below average levels can be observed across almost all reporting states.

**Figure 1:**
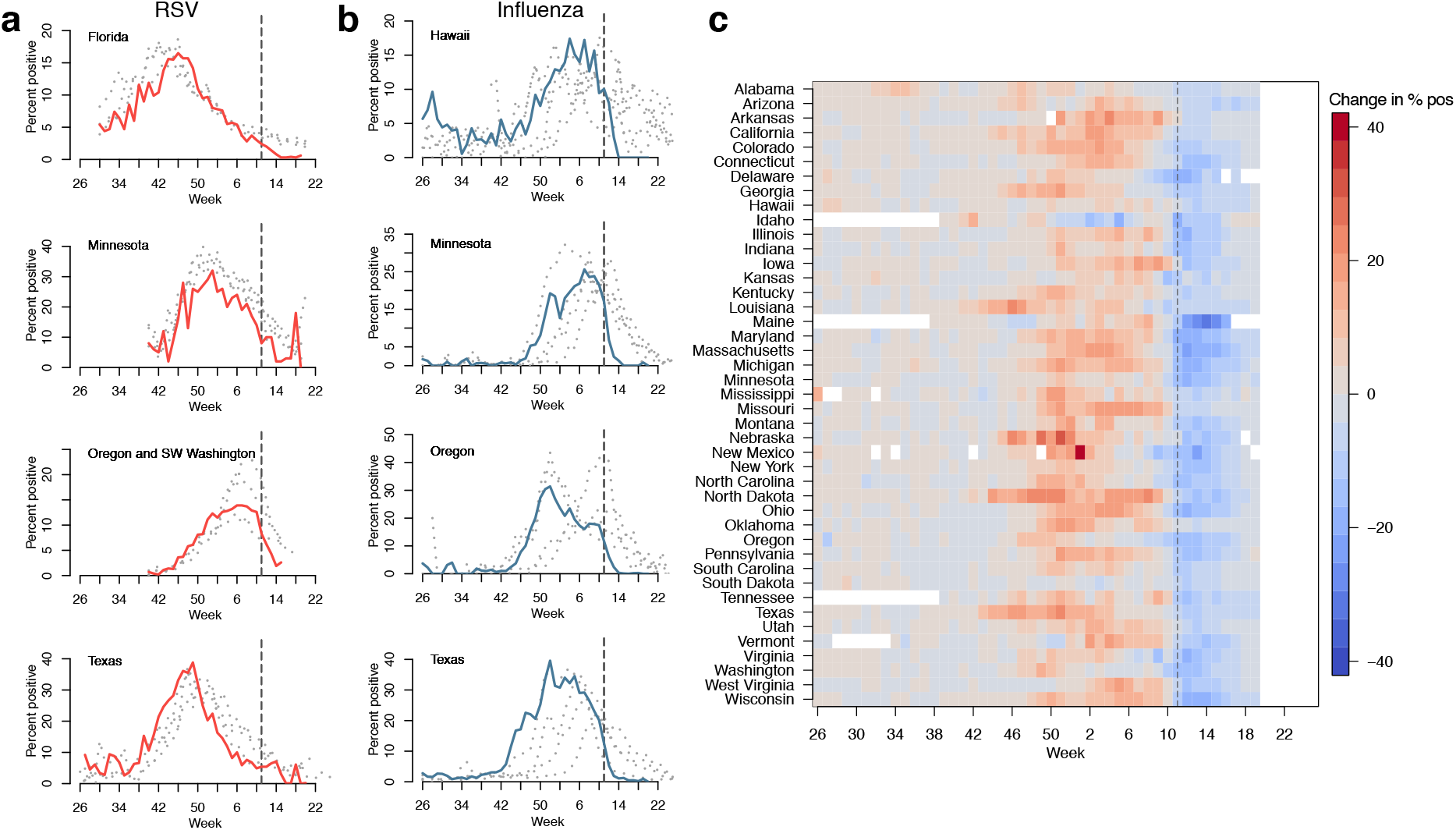
Reduction in RSV and Influenza cases since March 2020. The percent positive laboratory tests for a) RSV and b) Influenza across four US states. Data from 2020 are highlighted in red (RSV) and light blue (influenza). Data from previous seasons (2016-2019) are highlighted in grey. c) 2020 change relative to seasonal mean for influenza for all available US states (RSV surveillance data is only available for select states). Dashed lines show timing of the declaration of national emergency

To explore the possible implications of control (i.e. NPI) periods for the future dynamics of influenza and RSV we use epidemiological models and consider a range of possible scenarios for the length and intensity of control measures. Given the current uncertainty in the future course of the COVID-19 pandemic, and how responses might change over time, we cannot make precise predictions of future outcomes. For RSV, we use the time-series SIR model [26, 27], fitted to historic US case data described in previous work [11]. Specifically, we evaluate how NPI perturbations impact the epidemic limit cycles of RSV. We first consider a range of control period lengths and percent reduction in transmission based on Florida and Texas seasonality. Fig 2a,b show the impact of these varied controls on peak incidence (I/N), peak proportion susceptible (S/N) and timing of peak I/N.

**Figure 2:**
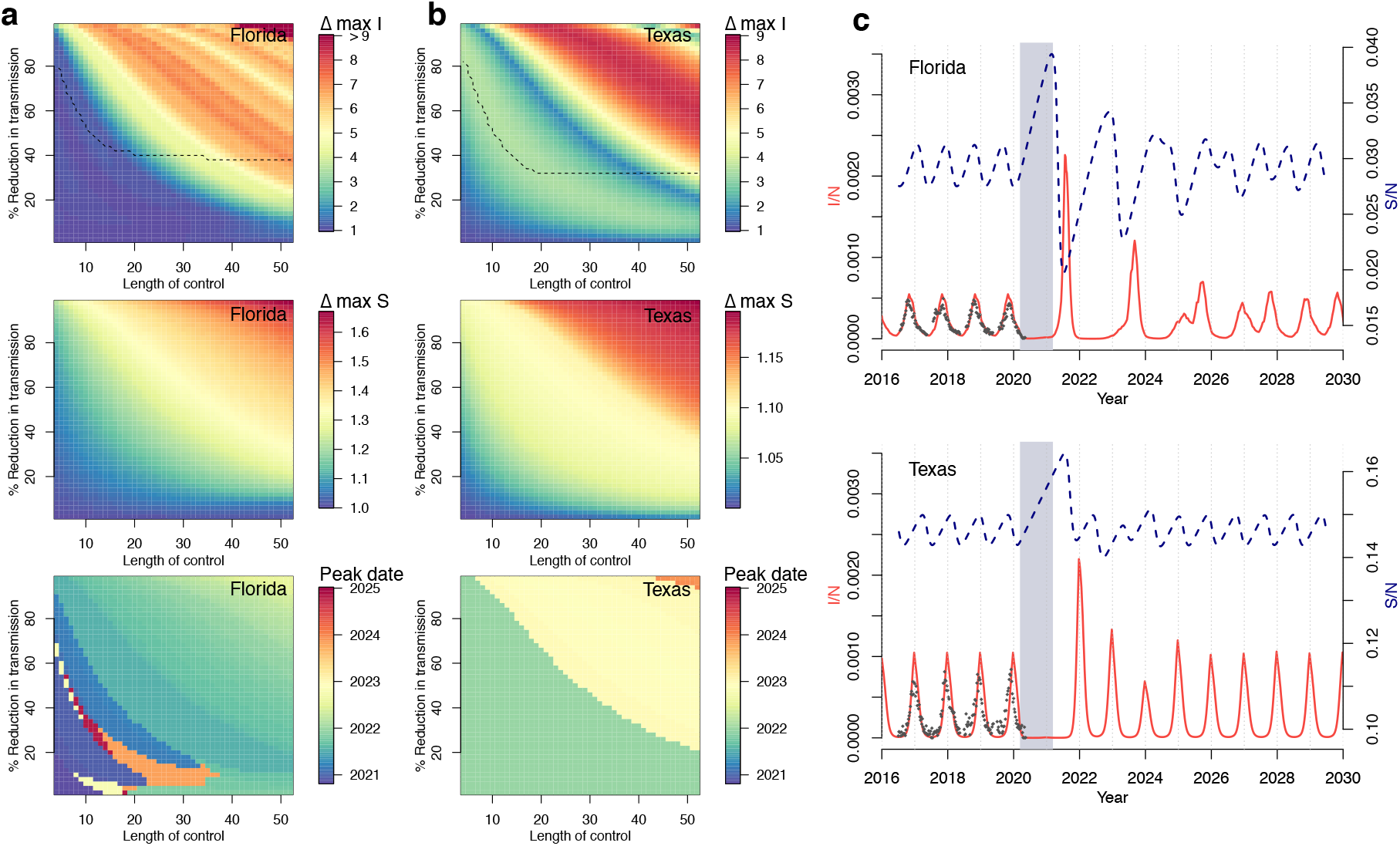
RSV simulations for Florida and Texas. Surface plots show the change in peak incidence per capita and peak susceptibility per capita, relative to pre-2020 maxima, for varied lengths of control (weeks) and % reduction in transmission. Black dashed line in the first plot row shows the region above which minimum incidence drops below 1, i.e. where local extinction is possible. The lower surface plot shows the timing of peak incidence in this period. Results for a) Florida and b) Texas are shown. Simulations of future RSV epidemics, assuming a control period of one year and a 20% reduction in transmission, are shown c) for Florida and Texas. Grey block represents the NPI period, red line is I/N and blue dashed line is S/N.

Major dynamic effects are caused by a buildup of susceptible individuals as NPIs reduce transmission. Longer controls, with a greater reduction in transmission, lead to a greater increase in susceptibility and larger resulting outbreaks. For Florida these outbreaks tend to occur in the summer months, but can occur throughout the year. For Texas, where seasonal transmission peaks in the winter, peak outbreaks occur only in the winter months, with the earliest outbreak in 2022.

We use RSV laboratory surveillance data for Florida and Texas to parameterize the actual reduction in transmission caused by SARS-CoV-2 NPIs. We find that a reduction in transmission of 20% is able to conservatively capture the decline in prevalence recently observed in the surveillance data (Fig. 2c, Fig. S3). Using this model parameterization, we run simulations with a control period of one year. Results from Florida and Texas, shown in Fig 2c, indicate an increased likelihood of severe RSV outbreaks after the control period has ended.

We then run simulations to investigate the potential impact of control measures on RSV for over 300 US counties and Mexican states using the time-series SIR model fitted to historic county-level case data (Fig 3) [11]. Counties with short time series (less than fives years of data) and sparse numbers of cases (under ten at peak) are removed. We compare the impact of two periods of control: lasting six months (Fig 3a,c) and lasting one year (Fig. 3b,d). Although the six month control period occurs outside the peak season of the virus, substantial RSV outbreaks are still projected as a lagged response to the SARS-CoV-2 NPIs. In general, the longer, one-year control period results in larger RSV outbreaks, yet complex interactions with seasonality arise. For New York county, the shorter control period results in a large outbreak in the following winter (2022), but the longer control period results in a more persistent but less intense outbreak. In contrast, a large RSV outbreak is observed in Miami after a year of control measures. In most cases, simulated dynamics eventually return to the pre-NPI attractor. For Boulder county, by contrast, control periods have complex interactions with the seasonal biennial epidemics of the disease. In these deterministic simulations, a longer control period in Boulder county causes the epidemic trajectory to shift to a different attractor (Fig 3e, Fig. S7). In general, the timing and size of future outbreaks will depend on the interaction between the dynamics of susceptibility and the seasonality of transmission.

**Figure 3:**
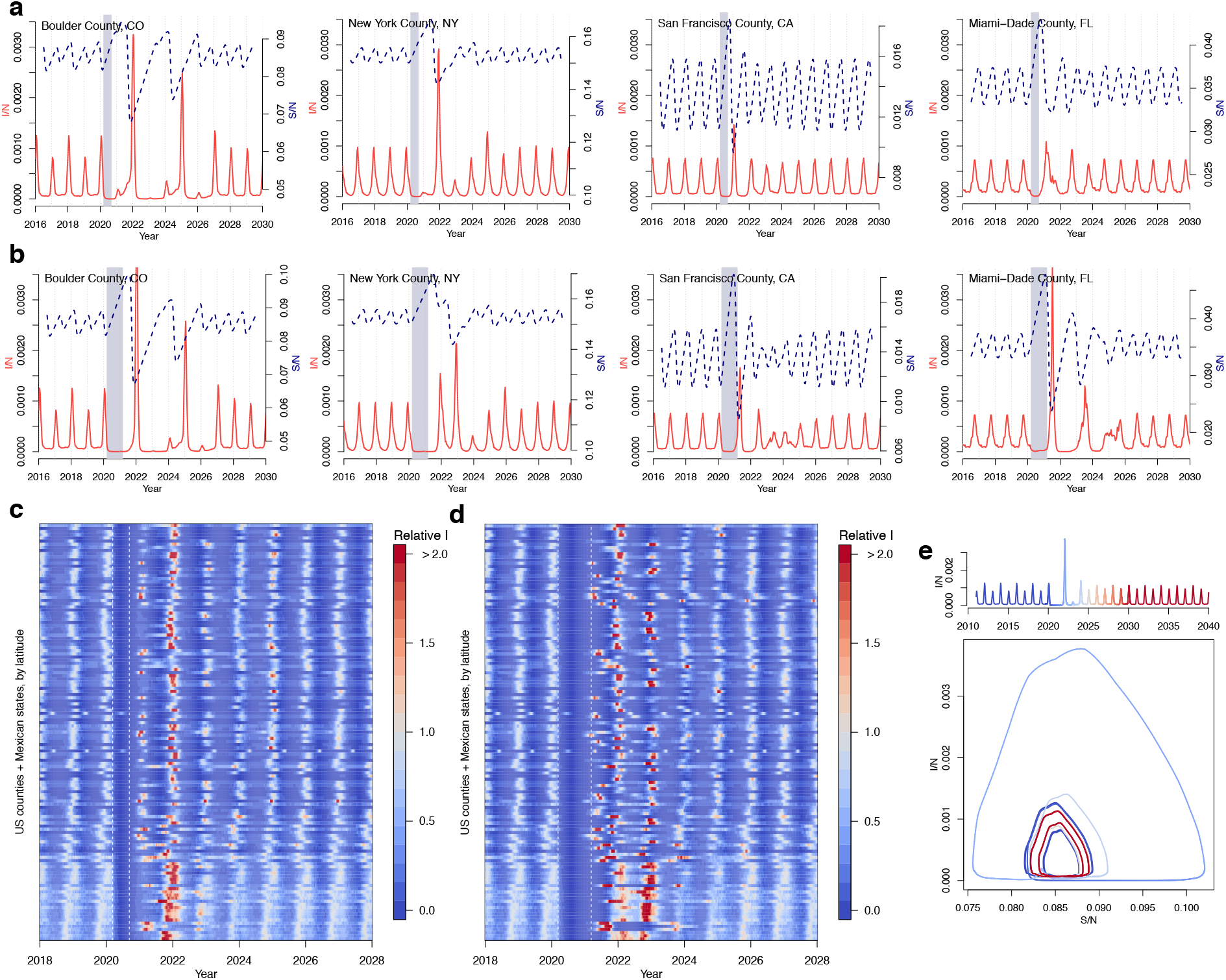
RSV simulations for US counties and Mexican states. Simulations for four US counties with either a) six month or b) one year of controls. Simulations for all US counties (with population *>* 500,000) and Mexican states in data with c) six month or d) one year control period, where max incidence prior to the control period is set to 1. e) SI phase plane plot for Boulder, Colorado showing epidemic trajectory with incidence time series above. The epidemic settles on a different attractor post-control.

Compared to RSV, influenza epidemics exhibit a less uniform seasonal pattern. Gradual evolution of the influenza virus’ antigenic sites (antigenic drift), means population susceptibility changes over time [21] and different subtypes may circulate each year with different levels of transmission [28]. In our preliminary analysis we therefore focus on the overall dynamics of susceptibility, ignoring year-to-year differences in circulating strains. We simulate influenza using a Susceptible-Infected-Recovered-Susceptible (SIRS) model, developed in previous analyses to explore influenza seasonality in the USA, where *R*_0_ varies between a maximum and minimum value driven by changes in absolute humidity [12, 13]. To capture the variability in transmission rates, we consider two scenarios: *R*_0*max*_ = 3 and *R*_0*max*_ = 2.2, based on the range of prior estimates [13, 28]. In both scenarios, *R*_0*min*_ = 1.2. We simulate the model using the climate of New York City.

Figure 4 shows the results using the influenza model under two control scenarios (six months and one year with a 20% reduction in transmission) and two transmission scenarios (high *R*_0_ and low *R*_0_). Six month controls have relatively little impact on influenza seasonality in New York in the high transmission scenario. In the equivalent lower transmission scenario, outbreaks after the NPI period are slightly elevated. In contrast, longer control periods provide more time for the susceptibility to build, resulting in an earlier influenza epidemic starting in the summer months. In the low transmission scenario, this is followed by a large outbreak in 2021. While these results suggest a more uncertain impact of NPI periods on future influenza outbreaks, dynamics will likely differ in locations with more persistent influenza cycles, such as the tropics [9].

**Figure 4:**
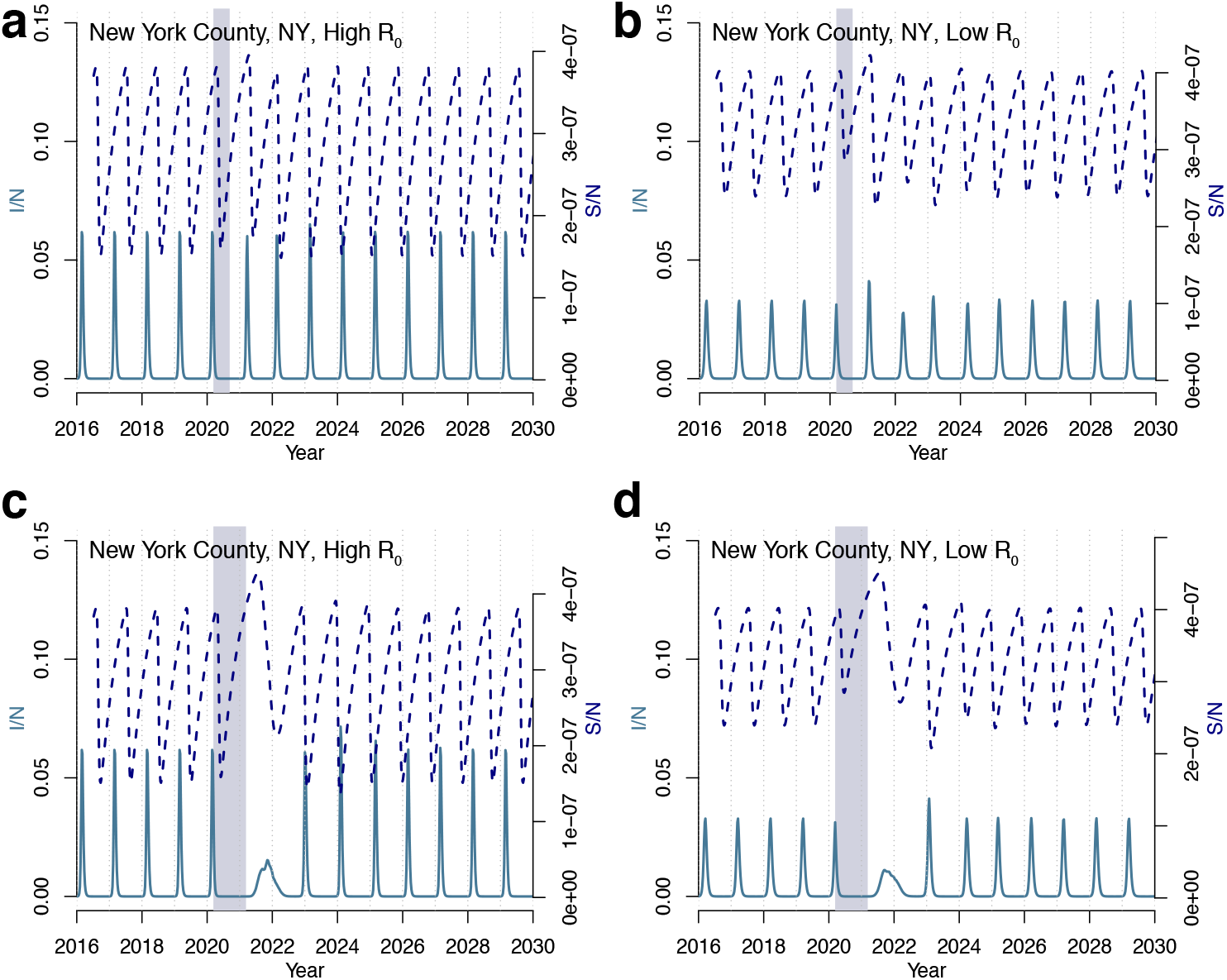
Influenza simulations for New York county. Simulations using a six month (a,b) and one year (c,d) control period for both high (*R*_0*max*_ = 3) (a,c) and low (*R*_0*max*_ = 2.2) (b,d) transmission rates.

Non-pharmaceutical interventions put in place to limit the spread of SARS-CoV-2 are already beginning to affect the transmission of other directly-transmitted, endemic diseases. Our results suggest that a build up of susceptibility during these control periods may result in large outbreaks in the coming years. Results for RSV in the USA suggest that these outbreaks may reach their peak in the winter of 2022. This finding appears robust even when we account for possible imported cases (results not shown). Following perturbation, RSV generally returns to the endemic attractor, but more complex behavior is possible (Fig. 3e).

Preliminary results for influenza suggest outbreaks may occur outside of the typical season, coinciding with the end of the control period. However, we do not address complex features of the influenza virus such as circulating subtypes or the implications of global NPIs for antigenic drift. The latter may prove significant, for example, if the evolution rate first declines with NPIs, then rebounds [29]. More broadly, our results suggest that healthcare systems may need to prepare for future outbreaks of non-COVID-19 infections, as NPIs are relaxed. These outbreaks may occur several years after initial NPIs were put into place.

There are several caveats to these results. First, we are at the early stages of understanding the implications of SARS-CoV-2 NPIs for endemic infections. In our model we used a fixed reduction in transmission however, this may not capture heterogeneities in NPIs across locations and over time. As more surveillance data becomes available, tracking further changes to endemic disease prevalence will be important. Serological surveys, currently used to measure exposure and potential immunity to SARS-CoV-2, could similarly be employed to monitor these polymicrobial responses [30, 31, 32]. Second, an influx of COVID-19 cases could artificially lower the percent positive test data we use to calibrate reductions in RSV and influenza transmission. For RSV this seems unlikely as the mean age of infection is much lower than COVID-19 and cases are unlikely to overlap. For influenza, this is more plausible, however, the sharp decrease observed across states right after the national emergency declaration suggests that it is unlikely to be primarily driven by this factor. Finally, interactions between the SARSCoV-2 virus and endemic viruses may be more complex than described here. Immunological relationships between viruses, both competitive and cooperative, may have broad scale implications for future infection dynamics [33]. The impact of NPIs on strain structure of RSV [34] is an important area for future work.

Finally, although we have primarily focused on the USA, outcomes may be more severe in southern hemisphere locations where NPI timing aligns with the peak season for seasonal wintertime diseases. Our results also illustrate the potential for COVID-19 NPI to impact the dynamics and persistence of a much wider range of infections. Increased surveillance, serological surveys and local modeling efforts will help determine the future dynamics and risk from these circulating infections.

## Methods

### Data

Recent (2016-2020) disease data based on laboratory results from either antigen or PCR tests for RSV are obtained from the corresponding government websites for each state: Florida (unspecified), Minnesota (antigen), Oregon (antigen and PCR), and Texas (antigen). Even though a few other states report RSV surveillance, we do not include them in our analysis as they do not provide information on RSV circulation from previous years. Some RSV data are extracted from the graphs of the state surveillance reports as raw values are unavailable. Influenza surveillance data are obtained from Centers for Disease Control and Prevention FluView Interactive. Historic RSV data (pre 2010) used to train the RSV model comes from hospitalizations data originally obtained from the State Inpatient Databases (SIDs) of the Healthcare Cost and Utilization Project (HCUP) maintained by the Agency for Healthcare Research and Quality (AHRQ). Population data for the US are obtained from publicly-available combined files of United States Census Bureau data available via the National Bureau of Economic Research. USA birth data are downloaded from the Centers for Disease Control. Transmission in the influenza model relies on specific humidity data taken from NASA’s Modern-Era Retrospective analysis for Research and Applications version 2 (MERRA-2) dataset.

### RSV seasonality

RSV epidemics exhibit distinct dynamic patterns driven by local climate [11, 25]. Locations in the USA with a large seasonal variation in specific humidity tend to experience biennial RSV dynamics where a large outbreak is followed by smaller outbreak. Locations with a moderate seasonal variation in specific humidity tend to experience annual outbreaks and locations with less seasonal variation in specific humidity tend to experience more persistent RSV outbreaks. In these latter locations, such as in Florida and southern Mexico, outbreaks tend to coincide with periods of elevated rainfall. RSV epidemic dynamics exhibit a stable limit cycle structure. For a few locations, co-existing attractors are possible, with perturbations to the system driving a shift to an alternate stable limit cycle (Fig. S7) [35]. Although there are two circulating RSV strains, strain-level data were not available for the USA, so our models add across strain structure [34].

### Models

We first calculate location-specific seasonal transmission rates using the time series Susceptible-Infected-Recovered model (TSIR), a discrete time adaptation of the SIR model [36, 26]. County-level transmission rates were calculated for a previous study [11]. The TSIR model describes the number infected and susceptible individuals as a set of difference equations. The number of susceptible individuals is given by:

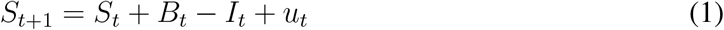

where *S*_*t*_ is the number of susceptible individuals, *I*_*t*_ is the number of infected individuals, *B*_*t*_ is births and *u*_*t*_ is additive noise, with *E*[*u*_*t*_] = 0. The time period *t* is the generation time for RSV, set at 1 week. The susceptible population can be rewritten as 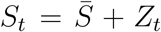 where 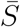 is the mean number of susceptible individuals in the population and *Z*_*t*_ is the unknown deviation from the mean number of susceptible individuals at each time step. Equation (1) is rewritten in terms of these deviations and iterated starting at *Z*_0_:

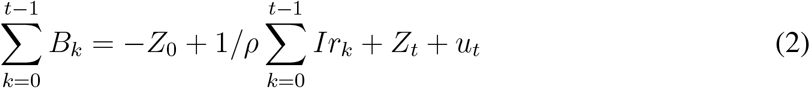

where *ρ* is the reporting rate which accounts for both under-reporting of RSV hospitalizations as well as infections that did not result in hospitalization and *Ir*_*k*_ is the reported incidence.

Using this formulation, it is shown that a linear regression of cumulative births on cumulative cases, gives *Z*_*t*_ as the residuals, assuming *u*_*t*_ is small. The inverse of the slope of the regression line provides an estimate of the reporting rate *ρ*. 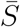 is calculated by defining the expected number of infected cases at each time step, *E*[*I*_*t*+1_], as:

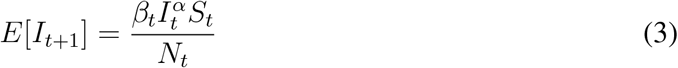

which is log-linearized as:

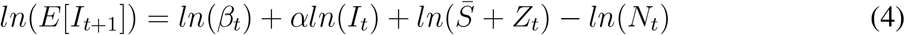

where *β*_*t*_ are biweekly factors that capture the seasonal trend in transmission rate and *α* is a constant that captures heterogeneities in mixing and the discretization of a continuous time process. We fix *α* at 0.97 to be consistent with prior studies [37, 11]. Equation 4 is fit using a Poisson regression with log link. The mean number of susceptible individuals, 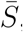, can then be estimated using marginal profile likelihoods from estimating equation (4), for a range of candidate values. Following [11], we add one to zero observations in the infected time series which represents continual low-level background transmission resulting in the lack of epidemic extinction we observe in the data. For fitting the TSIR we use the tsiR package [38]. When fitting to state-level data for Texas and Florida, we use a locally-varying spline regression for equation (2), which accounts for macro-scale changes in reporting over time.

We generate forward simulations using county seasonal transmission rates, *βt*, assuming a constant population and birth rate (based on average population and average birth rates from the historic time series). Model results are shown in terms of incidence per capita. The simulations are initially run for 40 years to remove transient dynamics. The control period is introduced to the model by lowering the seasonal transmission rates by a fixed proportion, starting on week 11 of 2020 (the week when a National Emergency was declared). For all simulations we lower the transmission by 20% unless otherwise specified.

The percentage reduction in transmission is estimated by comparing model simulations to laboratory RSV data from 2020 for Texas and Florida. State-level data from Minnesota are not used because the laboratory data does not capture the biennial cycles of incidence that exist in this state [11]. Other states do not have multiple years of available data to compare present reductions in prevalence. Lab test data are scaled to the model projection using the 2016-2020 mean i.e. 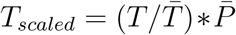 where *T* is the laboratory test data and *P* is the model projections. Simulations are run using reductions in transmission ranging from 0% to 90% in 10% intervals. For Florida, a 20% reduction in transmission, starting in week 11 (when the national emergency was declared), is found to be the best fit (Mean Absolute Error) reduction based on available data (Fig. S3). For Texas, both 10% reduction and 20% reduction give similarly good fits. Joint error is minimized using the 20% reduction rate.

For influenza we use a climate-driven Susceptible-Infected-Recovered-Susceptible model [13, 28, 14]. Antigenic drift of the influenza virus results in a seasonal return to susceptibility meaning TSIR methods are not appropriate for this infection. The model is described by a series of differential equations:

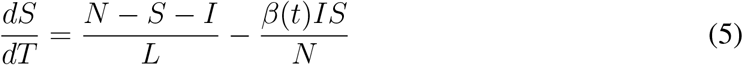

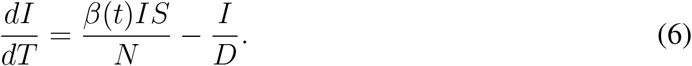

As before, *S* is the susceptible population, *I* is the number of infected individuals and *N* is the total population. *D*, the mean infectious period, is set a 4 days. *L*, the duration of immunity, is fixed at 40 weeks, allowing the influenza epidemic to recur each season. *β*(*t*) is the contact rate at time *t* and is related to the basic reproductive number by *R*_0_(*t*) = *β*(*t*)*D. R*_0_ is related to specific humidity *q*(*t*) using the equation:

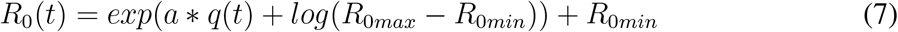

where *a* = −180, based on earlier findings [12, 13, 28]. *R*_0*min*_ is minimum reproductive number, fixed at 1.2, following [28]. *R*_0*max*_ is the maximum reproductive number. In Fig. 4, we use values of *R*_0*max*_ = 2.2 and *R*_0*max*_ = 3, based on plausible ranges observed in [13, 28].

## Data Availability

The US RSV hospitalization data are available from the Agency for Healthcare Research and Quality upon signing a data use agreement. Influenza surveillance data is publicly available from Centers for Disease Control and Prevention FluView Interactive. State-level RSV data is available from the public health departments of Minnesota, Florida, Oregon, Texas.

https://www.distributor.hcup-us.ahrq.gov/

https://www.health.state.mn.us/diseases/flu/stats/flustats19.pdf

http://www.floridahealth.gov/diseases-and-conditions/respiratory-syncytial-virus/_documents/2020-w19-rsv-summary.pdf

https://www.oregon.gov/oha/PH/DISEASESCONDITIONS/COMMUNICABLEDISEASE/DISEASESURVEILLANCEDATA/Documents/RSV_Oregon-2016-17.pdf

https://www.dshs.texas.gov/IDCU/disease/rsv/Data/2019-20/2019-20-RSV-by-DSHS-HSR-052020.pdf

## Funding

REB is supported by the Cooperative Institute for Modelling the Earth System (CIMES).

## Author contributions

Conceptualization: REB, SWP, CJEM, BTG; Data curation: REB, SWP, WY, GV; Formal analysis: REB, SWP; Methodology: REB, SWP, CJEM, BTG; Software: REB, SWP; Visualization: REB; Writing, original draft: REB; Writing, reviewing and editing: REB, SWP, WY, GAV, CJEM, BTG.

## Competing interests

The authors declare no competing interests.

## Notes

### Competing Interest Statement

The authors have declared no competing interest.

### Funding Statement

No external funding was received.

